# CSF estrogen, neuroinflammatory markers, and brain networks in middle-aged and older Black and White Women

**DOI:** 10.1101/2025.05.09.25327299

**Authors:** William T. Hu, Brittany Butts, Maria Misiura, Danielle D. Verble, Enid Swatson, Chloe Park, Jordan Watson, Bruno Hammerschlag, Ashima Nayyar, Naveen Korrapati, Lynn Marie Trotti, Karima Benameur, Laura M. Scorr, Michelle M. Mielke, Whitney Wharton

**Affiliations:** Cognitive Neurology and Alzheimer’s Disease, Rutgers-Robert Wood Johnson Medical School; Center for Healthy Aging Research, Rutgers Institute for Health, Rutgers Health, New Brunswick, New Jersey, USA; Nell Hodgson Woodruff School of Nursing and Emory University, Atlanta Georgia, USA; School of Medicine, Emory University, Atlanta Georgia, USA; Georgia State University, Atlanta, Georgia, USA; Department of Epidemiology and Prevention, Wake Forest University School of Medicine, Winston-Salem, North Carolina, USA

**Keywords:** Estrogen, health disparities, chemokine/cytokine, complement, hormone replacement therapy, MRI, Alzheimer’s disease

## Abstract

Neuroprotective properties of estrogen have poorly translated to reduced neurodegeneration in clinical trials of systemic estrogen replacement therapy. To more directly assess biological processes associated with brain estrogen (estrone, estradiol) levels, we recruited 81 women (42 non-white) and 28 men (13 non-white) for cerebrospinal fluid (CSF) proteomic and volumetric brain analysis. In these mostly post-menopausal women, we found low CSF estrogen levels to only modest correlate with their corresponding plasma levels. Aptamer-based proteomic analysis of CSF markers for inflammation, proteolysis, and DNA/RNA regulation revealed higher CSF estrogen to associate with changes involved in recruitment or activation of neutrophils, monocytes, and complement-related proteins in a race-dependent fashion. Parallel MRI analysis correlated higher CSF estrogen with smaller volumes of the brain somatosensory and posterior-medial networks without influence from cognition or neurodegeneration. These outcomes were only partially associated with plasma estrogens, reinforcing the need for improved CSF estrogen analysis to elucidate brain-specific effects.

## Introduction

Women and Black/African Americans (B/AA) face disproportionate risk for Alzheimer’s disease (AD) dementia, and the causes are complex.[1] In women, rates of brain atrophy associated with aging[2–4] or mild cognitive impairment (MCI)[5] differ from those in men. Conceptually, AD dementia disparities faced by women may result from hormone-related or hormone-independent processes related to biological sex as well as the social construction of gender.[6–8] The most frequently examined factor to account for AD dementia disparities between women and men has been exposure to sex hormones across the life span,[9, 10] as brain gray matter morphology, white matter hyperintensity, and functional connectivity undergo detectable changes during the menstrual cycle or menopause, and exposure to exogenous sex hormones.[11–16] While early observational studies showed reduced dementia risks with hormone replacement therapies (HRT),[17, 18] two large randomized controlled trials failed to demonstrate reduced dementia risks with HRT during early[19] or late[20] menopause. These negative prevention studies do not necessarily diminish the significance of endogenous sex hormones and their biological effects on the brain but do indicate the need for investigations beyond systemic (i.e. blood) circulating hormone levels.

In the brain, estrogen receptors are expressed in neurons as well as glia.[21, 22] Estrogen receptor binding leads to nuclear localization and transcription or membrane signaling independent of transcription.[23, 24] Ovariectomy in rats and non-human primates result in transcriptional changes in neurotransmitter receptors, neuropeptides, growth factors, and inflammatory mediators.[25, 26] More recently, human post-mortem brain multi-omics analysis in the Religious Order Study and Memory and Aging Project (ROS/MAP) showed overlap of genes associated with cognition and age at menopause as a trait.[27] However, there are key knowledge gaps between human blood-based and post-mortem neuropathologic studies to elucidate biochemical changes in the brain.[28–30] Proteins in the cerebrospinal fluid (CSF) better reflect brain-associated biological processes,[31, 32] yet we are not aware of any study having paired CSF sex hormone with CSF proteomic analysis.

For these reasons, we conducted a study to examine the relationships among sex, age, CSF sex hormones, and 1,075 CSF proteins related to inflammation, ubiquitination, cellular signaling, RNA processing, and other biological processes.[33] We recruited approximately equal numbers of B/AA and NHW women to examine these process for three reasons. First, older B/AA adults have 64% greater lifetime AD dementia risks than non-Hispanic White (NHW) adults, and understanding this difference can lead to identification of those at high risks regardless of race.[34] Second, we already demonstrated CSF biomarker differences between these two groups in tau-related biomarkers despite similar levels of beta-amyloid 1-42,[32, 35] AD-related cytokine profiles,[32] and blood-brain barrier integrity.[36] Finally, B/AA women also have earlier menarche,[37] higher menstrual E2 levels,[38] and earlier entry into menopause[39] than NHW women. B/AA race may thus be a surrogate marker for higher E2 levels, and reconciling these findings can elucidate race-independent factors underlying health disparities.

## RESULTS

### Men and B/AA post-menopausal women had higher plasma E1 and E2 levels

Between November 2020 and September 2024, 81 women and 28 men completed structured clinical, neuropsychological, brain MRI, plasma, and CSF analysis. Approximately three-quarters of the women (28/38 B/AA and 32/39 NHW, p=0.377) reported to be post-menopausal, and 12 (14.8%) had a history of hysterectomy only (n=7), oophorectomy only (n=1), or both (n=4). 10 women (12%) and 0 men were on HRT containing estrogen only (n=4), progestin only (n=3), or both (n=3).

Plasma and CSF sex hormones were measured using liquid chromatography-tandem mass spectrometry (LC-MS/MS) at the Mayo Clinic. Compared to men, post-menopausal women had lower plasma E1 (log_10_-transformed mean 1.21 vs. 1.45, p<0.001, Fig 1A) and E2 (log_10_-transformed mean 0.54 vs. 1.33, p<0.001, Fig 1B) but higher plasma SHBG levels (log_10_-transformed mean 1.59 vs. 1.40, p<0.001). Among post-menopausal women not on estrogen-containing hormone therapy (E+HRT), B/AA participants had greater plasma E1 and E2 levels than NHW participants. We did not detect this difference according to race in the smaller group of men.[40] In both women (Pearson’s correlation coefficient ρ=0.846, p<0.001)[41] and men (ρ =0.712, p<0.001), plasma E1 and E2 levels were highly correlated.

**Figure 1.**
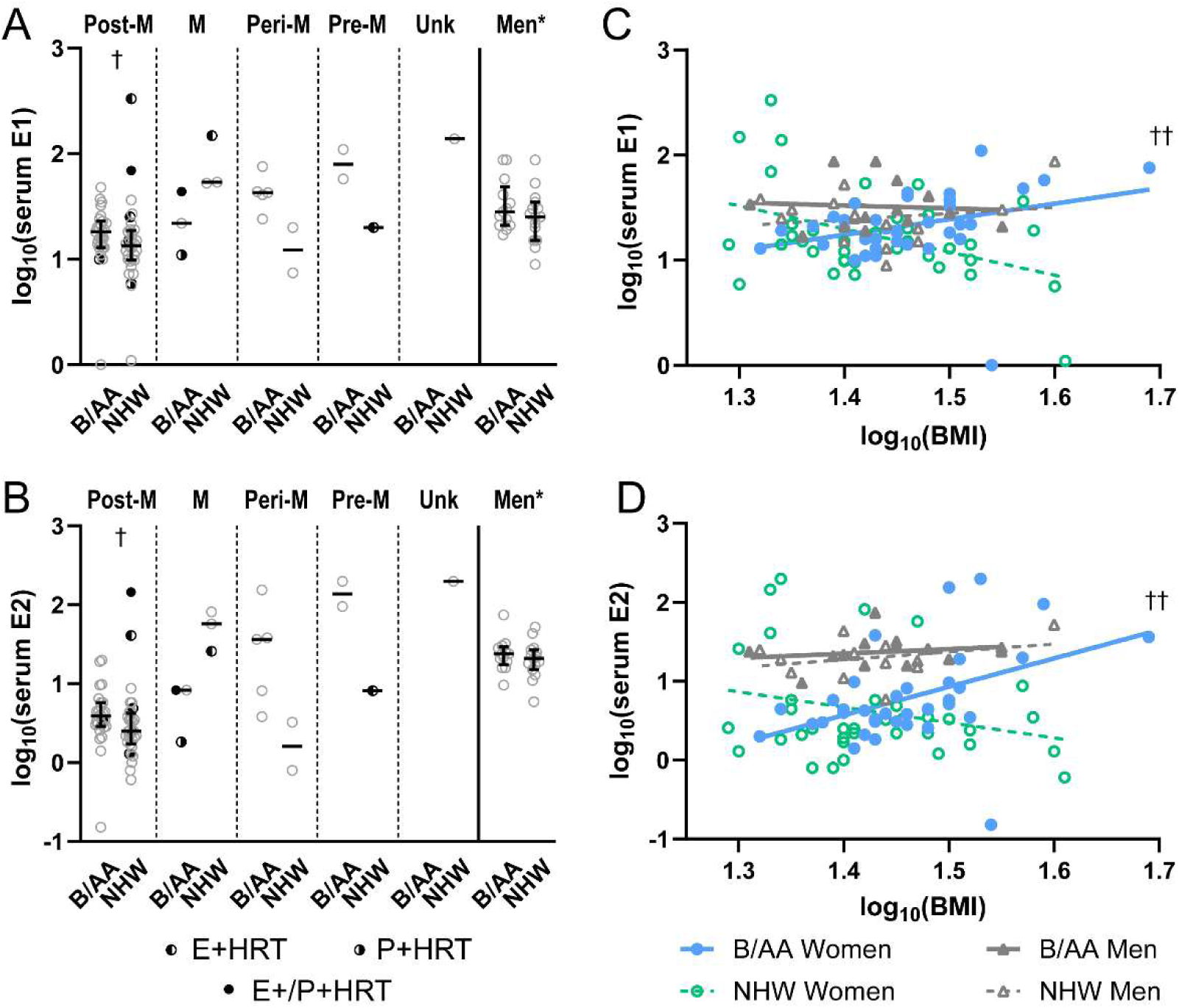
Plasma E1 and E2 levels according to sex, race, and BMI. B/AA and NHW participants were examined according to biological sex, self-reported race, and self-reported menopausal states in women (Post-M: post-menopausal; M: menopausal; Peri-M: peri-menopausal; Pre-M: pre-menopausal; Unk: unknown menopausal status; A & B) or BMI (C & D) for plasma E1 and E2 levels. For this analysis, women reporting other race (n=4) were excluded. Hormone replacement therapy (HRT) is shown to contain estrogen (E+), progestin (P+), or both (E+/P+). * Greater in men than all women combined, p<0.01; † Greater in B/AA than NHW women not on E+HRT, p<0.05; †† B/AA race X BMI interaction term, p<0.01.

### Race modifies the relationship between BMI and plasma estrogen levels in women, but not men

Seven of the 81 women were on estrogen-containing hormone replacement therapy (E+HRT), and they had higher plasma E1 (p=0.002), E2 (p=0.046), and SHBG (p=0.019) levels than those not on E+HRT. However, they also tended to be younger (58 vs. 63.5 yr, p=0.098) and have lower BMI (24.8 vs. 28.4 kg/m^2^, p=0.091). Linear regression analysis showed that higher plasma E1 levels was associated with lower BMI in NHW women (Fig 1C, 95% confidence interval of slope: -3.76, -0.64 for NHW vs. -0.061, +3.02 for B/AA women, p<0.005), and this difference persisted after adjusting for E+HRT and post-menopausal status (Table 2). A similar trend was observed for plasma E2 (Fig 1D, 95% confidence interval of slope: -4.271, +0.382 for NHW vs. +1.033, +6.169 for B/AA women, p=0.006), also persisting after adjusting for E+HRT and age (Table 2). No effect of race was seen on plasma E1 or E2 levels in men.

**Table 1.**
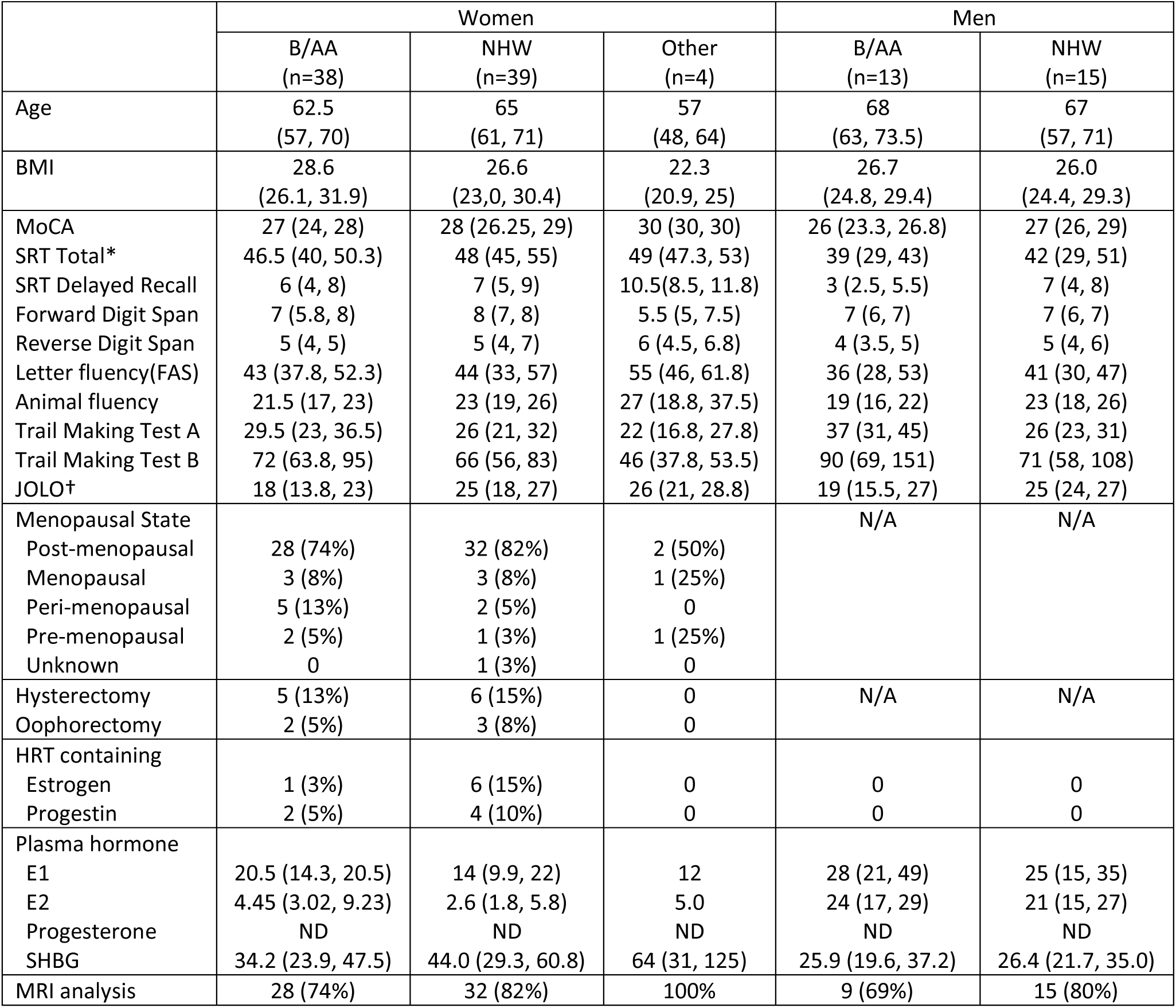
Demographic and sex hormone levels of participants. Median and interquartile ranges are shown for continuous variables (* greater in women than men, p=6.4 x 10^-4^; † greater in NHW than B/AA participants, p=3.5 x 10^-5^). MoCA: Montreal Cognitive Assessment; SRT: Buschke Selective Reminder Test; JOLO: Judgement of Line Orientation; HRT: hormone replacement therapy; E1: estrone; E2: estradiol; SHBG: sex hormone binding globulin; ND: not detected except in 3 women’s samples; N/A: not applicable to men.

**Table 2.**
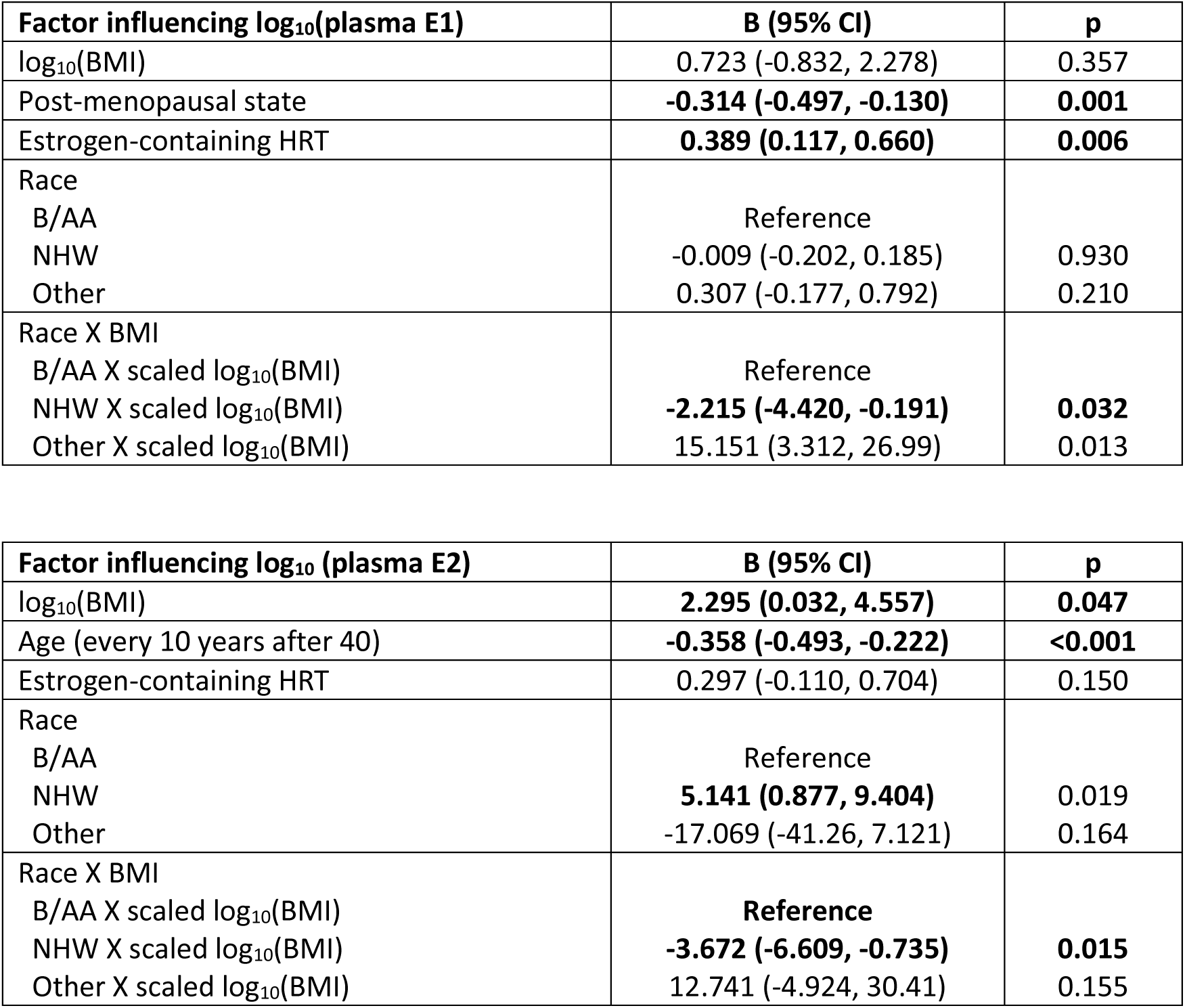
Factors associated with plasma E2 and E1 levels from linear regression.

**Table 3.**
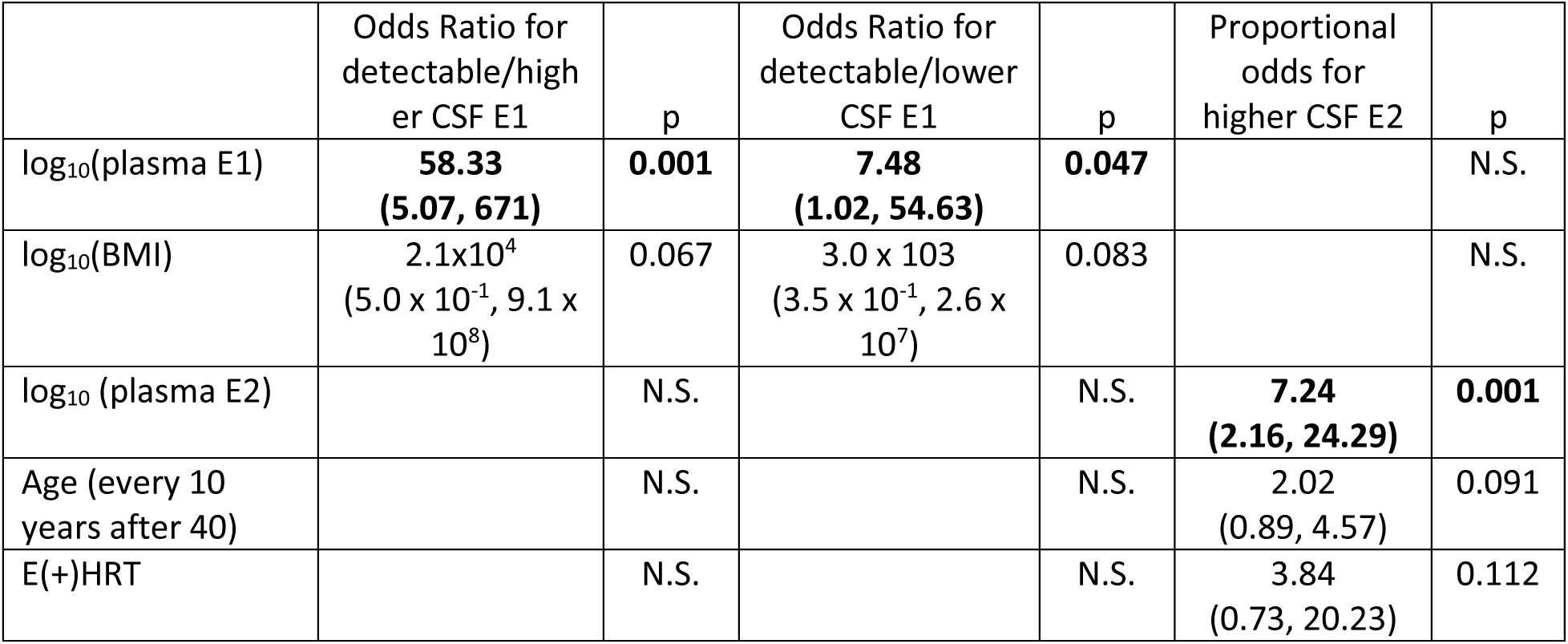
Association between CSF tertiles and plasma levels according to multi-nominal logistic regression analysis (for CSF E1, parallel assumption not met for ordinal regression) or ordinal regression analysis (for CSF E2). N.S.= not significant (factors with p≥0.150 at the model level are excluded).

### Effects of plasma estrogen, BMI, and age on CSF E1 and E2 levels

There was a moderate correlation between CSF and plasma levels of both E1 (ρ=0.600, p<0.001) and E2 (ρ=0.554, p<0.001). While race appeared to modify the relationship between plasma and CSF E2 levels for women (Fig 2A), this analysis was limited by the high number of samples having undetectable E1 or E2 levels (13 of 28 men and 56 of 81 women, including 3 of 7 women on E+HRT). We therefore used multinomial logistic regression (E1, due to failed parallel line assumption) or ordinal regression (E2) analysis to determine factors associated with higher CSF estrogen levels, categorized as undetectable, detectable/lower, detectable/higher (see Methods). High plasma levels were associated with higher CSF levels for both E1 and E2, while BMI correlated specifically with CSF E1 and age correlated specifically with CSF E2. After accounting for BMI and age, race did not appear to influence the relationship between plasma and CSF estrogen levels in this cohort (Table 2).

**Figure 2.**
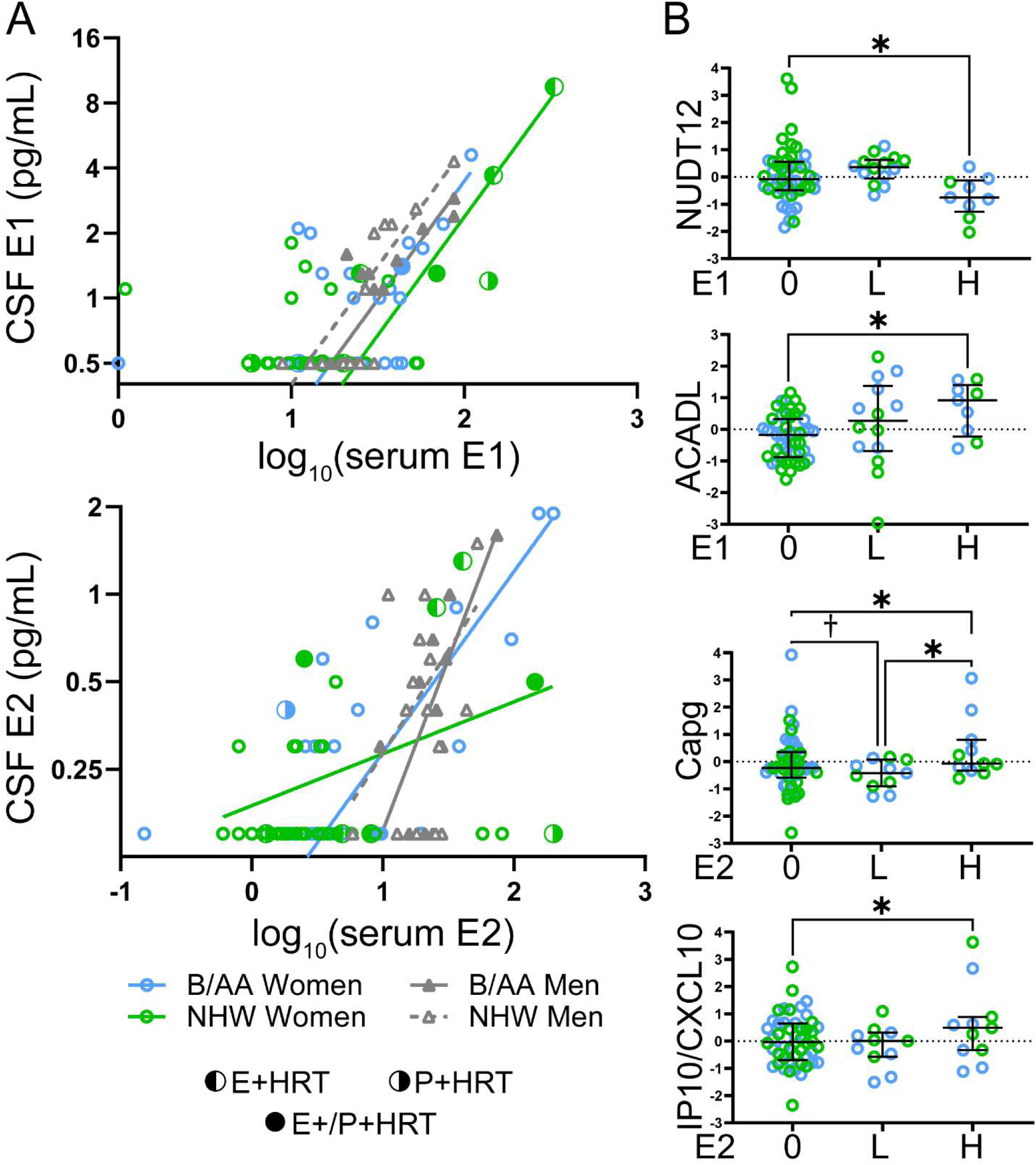
Relationship between plasma sex hormones, CSF sex hormones, and proteins related to age at menopause. CSF levels and E1 and E2 were detectable in fewer than half of the women (A), and were therefore analyzed as ordinal variables (0=undetectable, L=detectable/lower, H=detectable/higher). Hypothesis-based analysis using products of genes previously linked to age at menopause showed higher CSF E1 levels to associate with higher levels of nudix hydrolase 12 (NUDT12) and acyl-CoA dehydrogenase, long chain (ACADL); higher CSF E2 levels to associate with higher levels of capping actin protein, gelsolin-like (Capg) and interferon-induced protein 10 (IP10/CXCL10). * p<0.05 by multivariate multiple regression, † p=0.052.

### Effects of CSF estrogen, age, and HRT on CSF proteins

The association of BMI and older age with CSF estrogens in women is consistent with prior reports of non-gonadal estrogen sources including the brain. To identify brain processes potentially influenced by CSF estrogen levels, we analyzed proteomic correlates in 96 of 109 participants using aptamer-based SomaScan (SomaLogic, Boulder, CO; see Methods), which measured 1,075 proteins. We used two complementary approaches: (1) a hypothesis-driven analysis based on menopause-related genes from ROS/MAP, and (2) a data-driven approach using principal component analysis (PCA) of all measured proteins.

First, we performed a hypothesis-driven analysis based on 60 genes previously identified from post-mortem tissue in the ROS/MAP study as associated with age at menopause[27], which corresponded to 61 protein products in the SomaScan (Table S1.1). Multivariate multiple regression (MMR) analysis accounting for age, race, CSF E1 or E2 tertiles, HRT, and multiple hypothesis testing (see Methods) showed 21 of 61 CSF analytes to correlate with at least one of these factors at the model level. Models including CSF E2 tertiles showed greater influence from demographic factors than models including CSF E1 tertiles (Tables S1.2-S1.8). Four proteins were directly associated with CSF E1 and E2 levels (Fig 2B): NUDT12 (nudix hydrolase 12) and ACADL (mitochondrial long-chain specific acyl-CoA dehydrogenase) for E1, and CAPG (capping actin protein, gelsolin-like) and IP10/CXCL10 levels for CSF E2 (Tables S1.9-S1.12). Including an interaction term between race and CSF E1 levels identified two additional proteins with race-specific associations: CFB (complement factor B) and Aβ/APP (a SomaScan measure targeting but only modestly and inversely correlated with AD-related Aβ42).

While the above hypothesis-based approach isolated five identifiable CSF proteins which associated with CSF estrogen levels, it is difficult to extrapolate biological significance from these alone due to the limited targets examined.[42] We thus leveraged a systems biology approach to identify CSF proteomic features associated with CSF estrogen tertiles, age, race, HRT, and Montreal Cognitive Assessment (MoCA). Scaled levels of 1,075 CSF proteins were analyzed through principal component analysis (PCA) to generate 49 principal components (PCs, Table S2.1) representing protein groups or clusters which show co-variance across participants.[43] All PCs were then entered into an MMR to test for the effects of age, race, BMI, HRT, CSF estrogen tertile, and the interaction term between race and CSF estrogen tertile. For CSF E2, MMR showed eight PC scores to have model-level significance at p<0.05 (Table S2.2), with one associated with CSF E2 tertiles (PC43, p=0.006; Fig 3A, Fig S1A & B; Table S2.3) and two with race as well as the interaction between race and CSF E2 tertiles (PC19 & PC 42; Fig 3B, 3C, S1C-F; Tables S2.4 & S2.5). On the other hand, PC41 (Fig 3D & S1G-H; Table S2.6) and PC29 (Fig 3E & S1I-J; Table S2.7) were associated with the interaction between race and CSF E1 tertile, and PC29 was additionally associated with CSF E1 tertile (Table S2).

**Figure 3.**
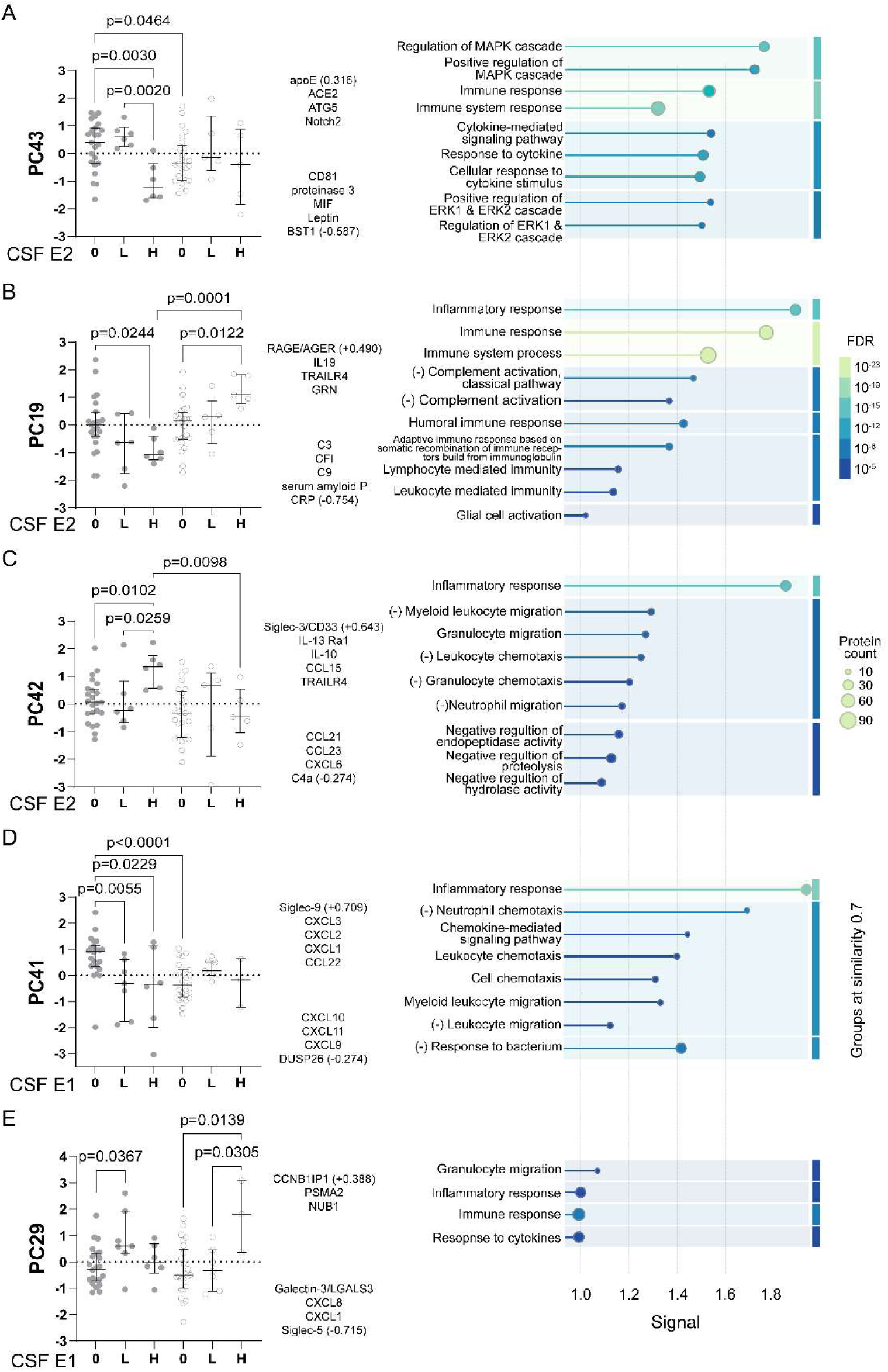
Relationship between CSF protein groups/modules and CSF estrogen levels. Principal components (PCs) derived from targeted CSF proteomics identified three to associate with CSF E2 (A-C) and two to associate with CSF E1 (D-E) tertiles (0=undetectable, L=detectable/lower, H=detectable/higher). Representative proteins positively or negatively loading onto each PC are shown (scores for the strongest loading protein in parentheses). Gene Ontology biological processes associated with each PC were identified according to number of proteins included in each process and enrichment signal, with stringent thresholds for enrichment signal (>=1.0) and false discovery rate (FDR) given the smaller gene sets from each PC entered into STRING-DB. Top generic process such as “inflammatory response” or “immune response” for each PC was included as reference for signal, protein count, and FDR.

**Figure 4.**
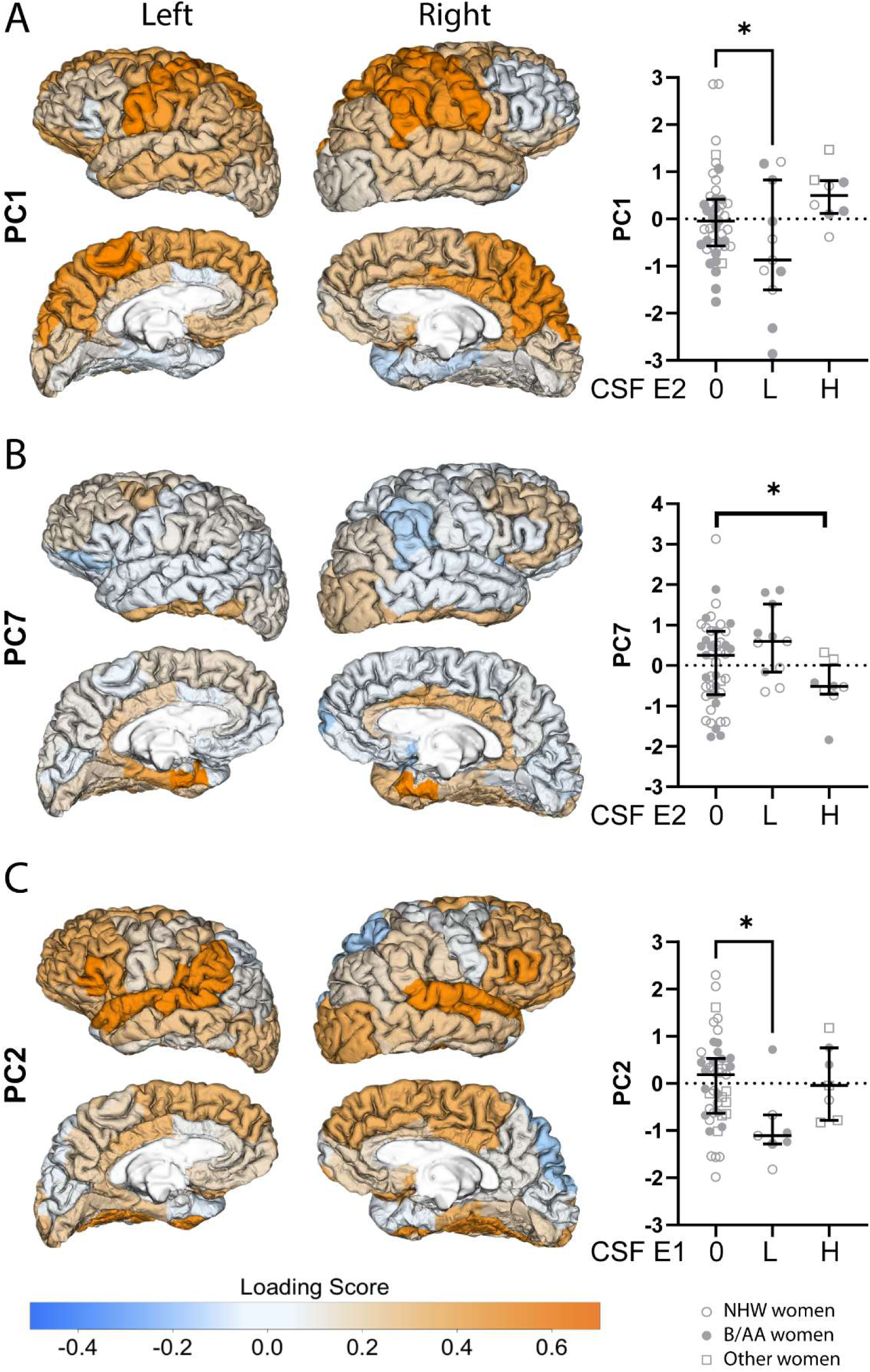
MRI correlates of CSF E2 and E1 levels. MRI principal components (PCs) corresponding to somatosensory (PC1, A) and posterior medial (PC7, B) networks differed according to CSF E2 tertiles, and MRI PC corresponding to default mode network (C) differed according to CSF E1 tertiles (0=undetectable, L=detectable/lower, H=detectable/higher).

Because of the disproportionate number of participants with undetectable CSF estrogen levels, we performed post-hoc analysis of relationships between plasma estrogen levels and these CSF PCs. When plasma E2 levels and E(+) HRT (two factors influencing CSF E2 levels) were included instead of CSF E2 tertiles, we also found PC43 to associate with plasma E2 (p=0.027) as well as E(+)HRT (p=0.039), PC19 with E(+)HRT (p=0.019), and PC42 with race (p=0.045). When plasma E1 levels were included in the analysis in place of CSF E1 tertiles, we found PC29 to associate with plasma E1 (p=0.077) but PC41 to associate with race (p=0.024) and age (p=0.003).

To better understand the biological functions represented by protein groups associated with CSF E2 and E1, we conducted pathway enrichment analysis using STRING-DB, a widely used resource for mapping protein interactions and enriched biological processes.[44] Given the intentional overrepresentation of inflammatory proteins in our SomaScan panel, we prioritized identifying more specific biological processes in Gene Ontology (GO) beyond general immune/inflammatory functions, which were only viewed as references. More GO biological processes were identified by STRING-DB to associate with CSF E2 than CSF E1 tertiles, but leukocyte proliferation/chemotaxis was represented in four out of the five PCs. At the same time, a closer examination of CSF analytes linked to these GO terms showed functional and/or structural specificity within each PC which informs potential mechanisms underlying these CSF protein groups/modules: analytes associated with *leukocyte proliferation* (e.g., bone marrow stromal cell antigen 1 [BST1], leptin [LEP], migration inhibitory factor [MIF], CD81) negatively loaded onto PC43; *CC chemokines* – C6-CC chemokines in particular, including CCL15 positively and CCL21/CCL23 negatively – loaded onto PC42;[45] CXC chemokines sharing receptor CXCR2 (CXCL1, CXCL2, CXCL3, associated with neutrophil chemotaxis[46–48]) positively and CXC chemokines sharing receptor CXCR3 (CXCL9 CXCL10, CXCL11, associated with monocyte chemotaxis) negatively loaded onto PC41.[49] Other unique processes associated with CSF E2 included positive loading of ERK1/2 cascade on PC43 (apoE, ACE2, Notch2; Fig 3A), positive loading of glia-related proteins on PC19 (RAGE/AGER, GRN, IL13), negative loading of complement regulation on PC19 (C2, C3, C5, C9, CFB, CFI, Fig 3B), and positive loading of markers for classical monocytes on PC42 (siglec-3/CD33, IL-13Ra1, IL-10).[50] These findings imply that, among women with undetectable CSF estrogen, B/AA participants show greater neuroinflammatory markers associated with neutrophil chemotaxis than NHW participants which diminish with higher CSF estrogen levels, but higher CSF estrogen levels are also accompanied by markers associated with complement activation in B/AA and monocyte/microglia activation in NHW participants.

### CSF E2 and E1 levels associated with large-scale brain networks

Finally, because estrogen can be secreted by or stored in cerebral structures, we examined whether CSF E2 levels associated with measurable differences in regional brain volumes among a subset of women who underwent MRI analysis (45, 11, and 8 women with undetectable, lower, and higher CSF E2 levels, respectively). Cortical volumes measured using FreeSurfer were first analyzed by PCA to identify eight PCs (Fig S2), with five PCs having significant loading from mostly symmetric bilateral structures (Table S3). Six PCs correspond to previously described brain networks including somatosensory (PC1, Fig 3A), default mode (PC2), limbic (PC3), visual (PC4), frontal pole auto-association,[51] and posterior-medial (PC7, Fig 3C) [52, 53] networks. The remaining two PCs correspond to the salience network (PC5, relatively more loading from left than right) and a network previously linked to movement initiation (PC8).[54] MMR of the eight cortical volume PC scores co-varying for total intracranial volume, age, race, BMI, HRT, and MoCA showed two brain volume PCs to associate with CSF E2 tertile (p=0.015 for Roy’s largest root, p=0.094 for Wilks’ lambda). Compared to participants with undetectable CSF E2 levels (Table S3), participants with detectable/lower CSF E2 levels had smaller PC1 (somatosensory network, -0.700 [-1.343, -0.057], p=0.033, Fig 3B) and participants with the detectable/higher CSF E2 levels had smaller PC7 (posterior medial, -0.839 [-1.614, -0.063], p=0.035, Fig 3D) volumes involving the entorhinal and parahippocampal regions. These are consistent with differences seen between men and women in the UK Biobank.[55] For CSF E1, participants with detectable but lower levels had smaller PC2 (default mode network, -0.867 [-1.532, -0.202], p=0.012) volumes. Thus, detectable CSF E2 or E1 levels were generally associated lower volumes of selective brain networks. However, this was not replicated when plasma estrogens were used in place of CSF estrogen, with PC1 volume instead having a positive correlation with plasma E2 (p=0.010) and E1 (0.016) levels in B/AA women.

## Discussion

The role of sex hormones on menopausal and post-menopausal women’s brain health remains controversial despite negative outcomes from the Women’s Health Initiative (WHI) and the Kronos Estrogen Prevention Study (KEEPS).[17–20, 56, 57] Here we prospectively measured E1 and E2 levels in a group of mostly post-menopausal B/AA and NHW women, and found only modest correlation between plasma and CSF levels. Using targeted CSF proteomics and volumetric MRI analysis, we also found higher CSF estrogen levels to correlate with neuroinflammatory markers suggesting a shift away from neutrophil chemotaxis towards monocyte chemotaxis according to race, and volumes differences in two networks previously linked to differ between men and women independent of race. We discuss these findings below.

Technical challenges in measuring estrogen levels are not new, and plasma measurements have more assay standardization than CSF measurements. Radioimmunoassay-based studies previously showed moderate correlation between CSF and plasma E2 across ages in women,[58–60] but a LC-MS/MS study showed stronger CSF-blood correlation for E1 than E2.[61] Because we observed race, age, and BMI to influence plasma and CSF estrogen levels, study participant characteristics very likely could lead to different effect sizes across studies. Notwithstanding these caveats, our data support positive relationships between plasma and CSF estrogen levels.[62]

While low CSF estrogen levels limit in-depth investigation into their source (circulation vs. brain) and impact, targeted CSF proteomics – favored over LC/MS-MS to investigate proteins of low abundance such as chemokines and cytokines – begins to identify protein groups/modules accompanying each CSF estrogen state. A main finding here is CSF estrogen levels’ preferential association with several inflammatory endophenotypes – some in race-dependent fashions. One example is the relative number of ligands for CXCR2 (CXCL1-3, involved in neutrophil chemotaxis)[46–48] to ligands for CXCR3 (CXCL9-11, involved in monocyte chemotaxis). Both groups/modules of ligands load onto PC41, but with associations in the opposite of their IFN-γ responses (positively for CXCL1-3, negatively for CXCL9-11).[63, 64] Greater PC41 scores in B/AA women than NHW women thus represent an extension of our previous finding of lower CSF CXCL9 levels in older B/AA than NHW participants,[32] and high PC41 scores in B/AA women with the lowest CSF E1 levels are conceptually linked to a shift from monocyte towards neutrophil chemotaxis. At the same time, higher CSF E2 levels are associated with higher complement protein levels in B/AA women but lower levels in NHW women, while higher CSF E2 levels also associate with a shift towards monocyte/microglial activation in NHW women. Thus, higher CSF E2 levels are associated with greater monocyte than neutrophil chemotaxis in both racial groups, even if baseline equilibrium between the two innate immune pathways differs according to race. A more neutrophil-biased CSF immune milieu in B/AA than NHW women generates new hypotheses relating to racial AD disparities, and should be thoroughly verified using cell-based techniques such as CSF single cell RNA sequencing.[33]

Aside from the possibility that estrogen mechanistically biases towards adaptive immunity,[65] it is similarly possible that estrogen alterations result from neuronal and glial alterations. Neurons can synthesize estrogen,[66] and brain injury is accompanied by increased aromatase expression as well as estrogen release by astrocytes.[67–69] Our brain volumetric analysis seems to support this explanation, showing smaller volumes in the somatosensory and posterior-medial networks for those with higher CSF E2 or E1 tertiles. While this appears consistent with the finding from WHI that women on conjugated estrogen HRT experienced greater brain atrophy,[70] estrogen-associated PCs loaded by neurodegenerative marker NfL (PC42) and A2 astrocyte marker S100A10 (PC19) showed changes in the opposite directions of neuronal injury in our study. What’s more, inclusion of MoCA to account for cognitive decline or neurodegeneration did not show a mediator effect between CSF estrogen tertiles and brain volume, and AD has been previously characterized by lower – not higher – brain[71] as well as circulating E2 levels.[72] Thus, relying on functional roles of estrogen in acute brain injury is insufficient to account for our observed association in our mostly cognitively normal cohort. Potentially more consistent with our observation are the observations that 1) estrogen can enhance the connectivity between the parahippocampal gyrus (part of the posterior-medial network enriched with estrogen receptors[73]) and other brain regions[74], and 2) increased connectivity can associate with lower regional gray matter volume via higher intracortical myelin content.[75] This alternate explanation should be tested in the future by examining white matter content and functional connectivity within the somatosensory and posterior-medial networks. If so, investigators should practice caution in interpreting gray matter volume-only findings especially in healthy older adults.

Our study has a number of limitations. The number of participants in each group and the proportion on HRT are small compared to WHI or KEEPS, but the in-depth CSF proteomic analysis conducted here is not feasible in large studies such as WHI or KEEPS. The technical sensitivity for CSF E2 and E1 is low compared to normative levels in this population, limiting our ability to fully model the effect of CSF estrogen on other CSF proteins on a continuous (rather than ordinal) scale. We did not incorporate CSF AD biomarkers in this analysis of participants with mostly normal cognition, although brain estrogen’s direct regulation of β-secretase [76, 77] and tau phosphorylation[78] potentially complicate interpretation of AD biomarker levels in this cohort. While we relied on bioinformatics to speculate on biological processes underling CSF protein changes, we did not provide cellular or other corollary to test these hypotheses. Even though we demonstrated an association between CSF estrogen and brain volumes, we could not identify the same relationship when plasma estrogen was used in place of CSF estrogen. Finally, we did not include genetic ancestry nor social determinants of health to further explore whether the race-associated modifications resulted from genetic or environmental differences, although the often strong collinearity between these factors calls for a well-balanced cohort in the future to address causal factors.

In conclusion, we used rigorous proteomic, MRI, statistical, and bioinformatics procedures to identify the impact of race on plasma estrogen levels, the relationship between plasma and CSF estrogen levels, and the association between CSF estrogen level and immune processes, but did not find race to influence network brain volumes. These results provide novel clues into estrogen-associated biological processes in the brain, reinforce the need to recruit diverse participants, and encourage caution when relying on plasma estrogen levels alone to examine sex hormones’ impact on the brain. Investigators should continue to explore high sensitivity assays to better detect CSF estrogen forms, and prospectively test the pathways hypothesized here towards future therapies to reduce sex- and race-related AD disparities.

### Experimental Model and Study Participant Details

#### Ethics Approval

This study was approved by the Emory University and Rutgers University Institutional Review Boards (IRB). Written informed consents were obtained from all participants.

#### Study Design and Recruitment

This parent study is longitudinal and on-going, and a cross-sectional design was used to characterize how estrogen levels correlate across biofluids (plasma, CSF), with CSF proteomics, and brain MRI volumes at baseline. Participants were recruited through health fairs, churches, Greek organizations, community presentations, Research Match, mailed research postcards, and from previous cohorts (PI Wharton) who agreed to future contact using Community Engaged Participatory Research (CBPR) principles. All research procedures take place at Emory University. Participants receive $300 total compensation.

Participants completed questionnaires, cognitive testing, fasting phlebotomy and lumbar puncture (LP), MRI analysis, vascular ultrasound, and overnight sleep monitoring using a single-lead EEG (Sleep Profiler). Participants were eligible if between the ages of 45-85; no/minimal memory complaints or diagnosis of MCI; MoCA ≥20 (n=81 for ≥26, n=17 between 23 and 25, n=11 for <23)[79]. Participants were excluded if there were contradiction for LP or MRI; residence in a skilled nursing facility; any significant systemic illness or unstable medical condition which could affect cognition, cause difficulty complying with the protocol, or consenting for study procedures; neurological disease which would influence cognition or CSF profiles (e.g., Parkinson’s disease, amyotrophic lateral sclerosis, multiple sclerosis, large territory stroke, traumatic brain injury); recent history of untreated major depression, substance use disorder; pregnant, nursing, or planning to get pregnant.

### Method Details

#### Questionnaires

Each participants completed questionnaires on medical and medication history, sleep (Epworth Sleepiness Scale), Patient Health Questionnaire 4, and Center for Epidemiologic Studies Depression Scale. In addition, they also completed a comprehensive questionnaire designed to characterize the sexual, reproductive and hormone-related history of middle-aged and older women, including current menopausal status and symptoms (if any), current or past hormone use (type, route), current use of aromatase inhibitors, and history of surgical hysterectomy or oophorectomy.

#### Neuropsychological Testing

Neuropsychological analysis was performed by experienced technicians to assess domain-specific cognitive functions, including verbal memory (Buschke Selective Reminding Test), attention/executive (Digit Span, Trail making test A&B, Symbol Digit Substitution Test), language (letter-guided fluency [FAS], Category Fluency-Animals), visuospatial (Judgement of Line Orientation) and overall (Montreal Cognitive Assessment) functions.

#### CSF and plasma collection

Twenty mL of CSF was collected via research LP using a 23.5 G non-traumatic Sprotte needle or a 22 G Quincke or Whitacre needle by a board-certified neurologist after overnight fasting. CSF was immediately (centrifuged at 4 °C for 10 min), aliquoted, labeled, and frozen until analysis. Approximately 45 mL of blood was collected on the same day in K2-EDTA tubes (for plasma) and SST tubes (for serum). For plasma, blood was centrifuged at 1000 g and 4 °C for 15 min before aliquoting, labeling, and freezing at -80°C until analysis.

#### Plasma and CSF sex hormone analysis

Plasma and CSF levels of E1, E2, progestin, and SHBG were measured using mass spectrometry at Mayo Clinic in a manner consistent with CLIA requirements.

For estrogen, E2 and E1 were extracted from 0.5 mL of biofluid (plasma or CSF) using methylene chloride, and Deuterated 17β-estradiol-d5 and estrone-d4 were added to each sample before the liquid extraction as internal standards. After derivatization with dansyl chloride, sample extracts underwent high-pressure liquid chromatography (HPLC) (Agilent Technologies 1290 Infinity UPLC ) and then LC-MS/MS (Agilent Technologies 6490 Triple Quadrupole; ESI interface, operated in the multiple-reaction monitoring positive mode). Concentrations were derived on a nine-point standard curve (range of 0–100 pg/mL).

For progesterone, levels were measured using a competitive electrochemiluminescence assay (Cobas e411, Roche Diagnostics) following manufacturer’s protoocl. The lowest level of quantitation (LLOQ) was 1.00 pg/mL for E1, 0.30 pg/mL for E2, 0.20 pg/mL for progesterone, and 5 pg/mL for SHBG. Concentrations below these levels were substituted with LLOQ/2 for subsequent analysis.

Concentrations were determined via a instrument-generated calibration curve.For SHBG, a microparticle-based immunoassay (Cobas e411, Roche Diagnostics) was used following manufacturer’s protocol.

#### CSF inflammatory protein analysis

A custom CSF targeted proteomic array (1,500 assays, with 33 targeting duplicate proteins to identify differences between assay versions) was developed using proteins associated with inflammation, transcriptional regulation, ubiquitination, and organellar functions (e.g., lysosome) from the greater SomaLogic CSF panel (Table S2). Frozen CSF samples were shipped from Emory University over-night on dry ice to Rutgers, randomized, and then sent to SomaLogic. Because CSF protein levels were influenced by freeze-thawing cycles during internal pilot experiments conducted by Rutgers and SomaLogic, samples were only thawed at time of analysis to minimize this effect.

In addition to participant samples, five pairs of adjacent CSF aliquots were sent to undergo the same SomaScan. For each analyte, quality control (QC) steps included assessment for quantitation and intermediate precision. Limit of quantitation (LOQ) was determined using a conservative threshold based on 10 blank samples (mean + 2 SD). Out of 1,467 analytes, 365 were below LOQ and excluded from subsequent analysis, while 27 had coefficients of variation (CV) ≥20%. The final SomaScan panel for analysis included 734 proteins with CV<10% and 341 with 10≤CV<20%.

#### Neuroimaging

A subset of participants (64 women and 24 men) underwent a 1 hour structural MRI analysis at the Emory Center for Systems Imaging. Sex distribution was similar between those with and without MRI (p=0.261). Among women, there was no difference in proportions reporting B/AA race (p=0.600), menopause (p=0.467), history of hysterectomy (p=1.000), history of oophorectomy (p=1.000), current E(+)HRT (p=0.613), and current P(+)HRT (p=0.588). MRIs were conducted mostly at the same visit as blood and CSF were collected, but no more than three months after. High-resolution anatomical images were acquired using a T1-weighted magnetization-prepared rapid gradient-echo (MPRAGE) sequence.

Imaging parameters included a repetition time (TR) of 2300 ms, an echo time (TE) of 2.95 ms, and an inversion time (TI) of 900 ms. The flip angle was set to 9 degrees. Images were obtained with a field of view (FOV) of 256 × 256 mm and a matrix size of 256 × 256, yielding a voxel size of 1.0 × 1.0 × 1.2 mm³. The slice thickness was 1.2 mm, with a total of 160 slices acquired. The acquisition time for this sequence was approximately 5 minutes. This sequence provided high-resolution anatomical images optimized for gray and white matter differentiation, suitable for volumetric and structural analyses.

Cortical reconstruction and volumetric segmentation were performed with the FreeSurfer image analysis suite (version 7.4.1), which is documented and freely available for download online (http://surfer.nmr.mgh.harvard.edu/). Volumetric estimates for regions of interest were derived based on the Automated Anatomical Labeling (AAL) atlas.[80]

### Quantitation and Statistical Analysis

#### Missing Data

Participants with missing plasma or CSF E1 and E2 data were excluded from the baseline and subsequent analyses. Only participants with CSF SomaScan results (96/109, 88%) were included for correlation with CSF proteomics.

#### Analysis of baseline characteristics

All demographic and clinical data were analyzed using IBM SPSS 28 (Aramonk, NY). For baseline comparisons, Student’s T-test or analysis of covariance (ANOVA) were used for continuous variables, while Chi-squared tests were used for categorical variables. For MoCA, the negatively skewed data were transformed by reflection, log_10_ transformation, and then reflection again for ease of interpretation.

#### Plasma and CSF sex hormone levels

Plasma E1 and E2 levels were log_10_-transformed before analysis due to non-normal distribution. Because significant numbers of participants had undetectable CSF E1 or E2 levels, each CSF sex hormone was analyzed as undetectable, detectable/lower, and detectable/higher across gender and racial backgrounds. For CSF E1, the three groups’ concentrations were < 1.0 pg/mL (n=70), 1.0-1.4 pg/mL (n=20), and 1.5-9.5 pg/mL (n=20). For E2, the three groups’ concentrations were < 0.30 pg/mL (n=65), 0.3-0.4 pg/mL (n=21), and 0.5-1.9 pg/mL (n=24).

#### CSF inflammatory protein analysis

After QC steps above, 1,075 CSF analyte levels underwent standardization and outlier detection across both sexes. PCA was then conducted in women only to identify CSF proteomic PCs.[33, 43] Briefly, each individual CSF protein was first examined to assess for normal distribution across the sample cohort using Kolmogorov-Smirnov Test, and CSF proteins which did not have normal distribution were log_10_-transformed. After log_10_-transformation, all CSF analytes were standardized, and outliers (≥4.0 or ≤-4.0) were imputed in SPSS using K-Nearest Neighbors (n=5). PCA was conducted using co-variance matrix and Varimax rotation, and the optimal number of PCs was selected based on 90% variance to maximize interpretability given the overall sample size as an eigen-value threshold gave too many PCs and there was no clear “elbow”.

#### Neuroimaging

Volumes of 34 cortical regions on each side were standardized before PCA (co-variance matrix, Varimax rotation). Elbow rule and total variance accounted by PCs were used on conjunction to determine eight PCs as the optimal solution. While left and right brain structures were allowed to independently undergo PCA, all PCs had loading from bilateral brain structures.

#### Association between estrogen levels and proteomic/MRI features

MMR analysis was performed with CSF or structural MRI PCs in women only. For each, CSF E2 level (undetectable, lower, higher), race, menopausal status, HRT, age, and MoCA were entered as independent variables, with multiple hypothesis testing adjusted at the MMR level. Total intracranial volume was also entered as an independent variable for structural MRI PC analysis. Only corrected models having omnibus significance (p<0.05) were examined in post-hoc analysis for directions of change. For CSF proteomic PCs, a race x CSF E2 tertile interaction term was entered as this improved the overall models. Omnibus parameters (Wilks lambda, Roy’s Largest Root) were first examined for the independent variables, and overall models with p<0.05 were then examined for between-subjects effects. Top positive and negative loading proteins were entered into STRING-DB using GO Biological Processes to identify pathways associated with changes in PC scores.[44] A similarity score of 0.7 was used to group GO processes sharing significant gene/protein products, and the top generic process such as “inflammatory response” or “immune response” for each PC was included as reference for signal, protein count, and FDR. The same was then repeated for CSF E1 tertiles, log_10_(plasma E2), and log_10_(plasma E1). For volumetric MRI PCs, only the interaction term between race and plasma estrogen levels achieved omnibus significance.

## Data Availability

The data that support the findings of this study are available from the corresponding author upon reasonable request.

## Author contributions

WTH and WW were responsible for conception/design the study; WTH, BB, MM, DDV, CP, JW, BH, AN, NK, LMT, KB, LMS, HZ, WW were responsible for acquisition, analysis, and interpretation of data; WTH, MM, DDV, NK, WW were responsible for drafting the work, and BB, ES, CP, JW, BH, AN, LMT, KB, LMS, HZ were responsible for critical revision of the work for important intellectual content. All authors have given final approval for the version submitted, and agree to be accountable for all aspects of the work.

