## Supplementary Figures for "CSF estrogen, neuroinflammatory markers, and brain networks in middle-aged and older Black and White Women"

**Supplementary Information**

**Table S1.** Targeted analysis of CSF SomaScan analytes overlapping with gene products linked to age at menopause (natural or surgical) in ROS/MAP. Among 2,685 brain-derived genes associated with age at menopause, 645 were associated with both natural and surgical menopause (1.1). Among these, protein products of 94 were measured in CSF SomaScan but 34 products failed quality control (QC) during analysis. Multivariate multiple regression analysis using 61 CSF proteins corresponding to the 60 remaining genes to examine differences according to E2 (1.2) and E1 (1.3) tertiles revealed age, race, and P(+)HRT to influence CSF SomaScan protein levels (1.4-1.8). In addition, CSF E2 tertiles were associated with Capg (1.9) and IP10/CXCL10 (1.10) levels, while CSF E1 tertiles were associated with ACADL levels (1.11).

**Table S2.** CSF SomaScan analytes associated with CSF E2 and E1 tertiles. 1075 SomaScan analytes first underwent principal component (PC) analysis to derive 49 orthogonal PCs (S2.1) and then multivariate multiple regression to account for multiple hypothesis testing (S2.2). Three PCs (S2.3, 2.4, 2.5) were associated with CSF E2 tertiles and two PCs (S2.6, S2.7) were associated with CSF E1 tertiles.

**Table S3.** Regional brain volumes linked through large scale networks associated with CSF E2 tertiles. 68 FreeSurfer brain region volumes first underwent principal component (PC) analysis to derive 8 orthogonal PCs (S3.1). Multiple regression analysis to account for multiple hypothesis testing only revealed two PCs (S3.2, S3.3) to associate with CSF E2 tertiles (after adjusting for age and total intracranial volume) and PC2 to associate with CSF E1 tertiles (S3.4).

**Figure S1.** CSF SomaScan PCs associated with CSF E2 (A, B, C) or E1 (D, E) levels and representative analytes loading onto these PCs. PCs were identified using multivariate multiple regression analysis, but p-values from pair-wise Mann-Whitney U-tests are shown to represent within and between race effects according to CSF estrogen levels. No post-hoc comparisons were made for SomaScan analytes loading onto these PCs, with loading scores (positive or negative) in parentheses.

**Figure S2.** PCA of regional brain volumes derived from FreeSurfer reproduce large scale brain networks including somatosensory (PC1), default mode (PC2), limbic (PC3), visual (PC4), left-predominant salience (PC5), frontal pole-associated (PC6), posterior-medial (PC7), and movement initiation-related networks (PC8).

**Figure S1.**

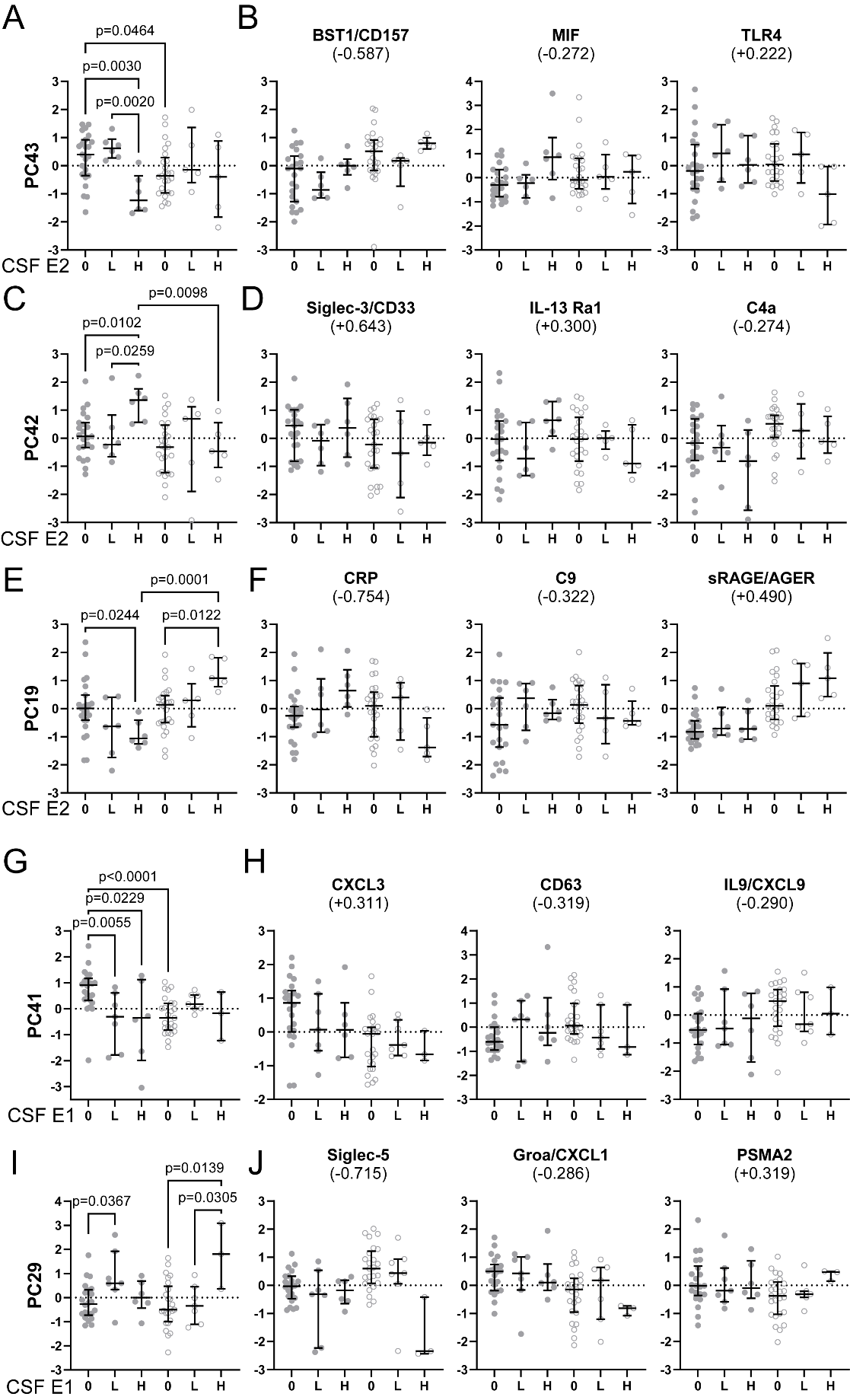

**Figure S2.**

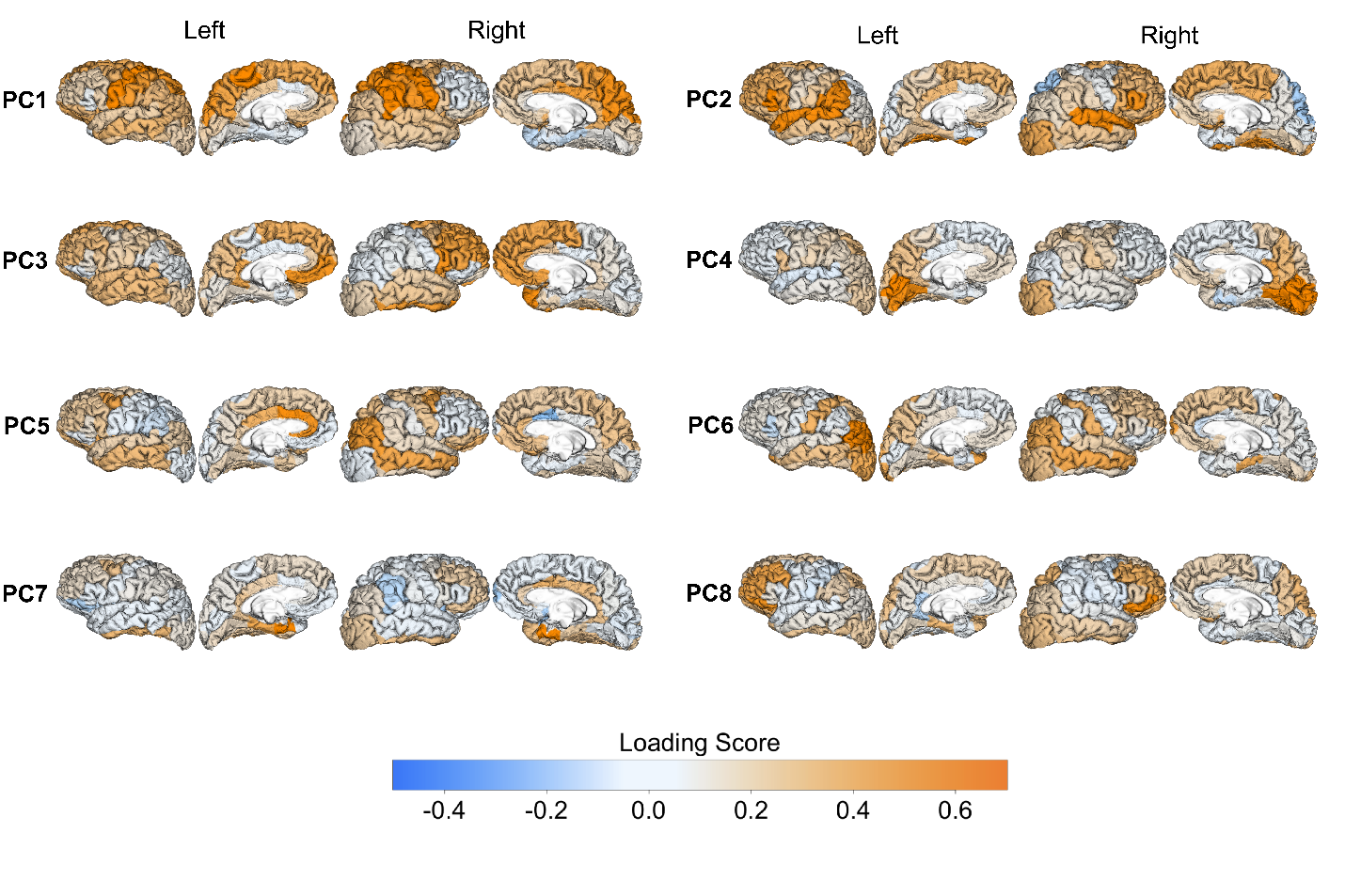
